# RNA sequencing resolves cryptic pathogenic variants in mitochondrial disease

**DOI:** 10.64898/2026.02.23.26345976

**Authors:** Zhimei Liu, Xin Duan, Fatemeh Peymani, Jia Wang, Chengjia Bao, Chaolong Xu, Ying Zou, Zixuan Zhang, Yunxi Zhang, Tongyue Li, Martin Pavlov, Junling Wang, Minhan Song, Tianyu Song, Xiaodi Han, Mingxi Sun, Danmin Shen, Ruoyu Duan, Huafang Jiang, Manting Xu, Holger Prokisch, Fang Fang

## Abstract

**Background:** Mitochondrial diseases are the most common inherited metabolic disorders, characterized by pronounced clinical and genetic heterogeneity that complicates molecular diagnosis. Although DNA-based sequencing approaches have become standard in genetic testing, up to half of patients remain without a definitive diagnosis. RNA sequencing (RNA-seq) provides a complementary layer of evidence by revealing functional consequences of genetic variation, thereby improving diagnostic yield.

**Methods:** We performed RNA-seq on skin fibroblasts from 140 pediatric patients with suspected mitochondrial disease who remained genetically undiagnosed after whole exome sequencing (WES). Aberrant RNA expression and splicing were identified using the detection of RNA outliers pipeline (DROP). Based on WES findings, patients were stratified into a candidate group (n=28), in which RNA-seq evaluated the pathogenicity of WES-identified variants of uncertain significance and an unsolved group (n=112), in which RNA-seq was used to pinpoint candidate genes. In six cases where RNA-seq identified the aberrant RNA-event but WES did not detect the causative variants, whole genome sequencing (WGS) was performed.

**Results:** Integrative RNA-seq, WES, and WGS analysis resulted in a genetic diagnosis in 25% of patients overall (20/28 [71%] in the candidate group; 15/112 [13%] in the unsolved group). Aberrant splicing explained most candidate-group diagnoses, including variants misclassified by in silico predictors such as SpliceAI. Fourteen percent of protein-truncating variants predicted to undergo nonsense-mediated decay (NMD) escaped degradation, highlighting the functional limits of current predictions. The variants identified in the unsolved cohort included synonymous, missense, deep intronic, near-splice-site variants, and large deletions. The most frequent amongst them was a recurrent synonymous East Asian founder mutation in *ECHS1*, accounting for seven cases. Interestingly, across 231 pathogenic variants associated with aberrant RNA phenotypes compiled from this study and prior reports, half were non-coding and half were coding variants.

**Conclusion:** RNA-seq substantially enhances molecular diagnosis in mitochondrial disease by exposing cryptic splicing, regulatory, and NMD-escape events invisible to DNA sequencing alone. These data advocate transcriptome analysis as an essential component of comprehensive genomic diagnostics in neuro-metabolic disease.

**Significance Statement:** Mitochondrial diseases remain among the most challenging inherited metabolic disorders to diagnose, with nearly half of patients unresolved despite advanced DNA sequencing. By integrating transcriptome profiling into the diagnostic workflow, this study demonstrates that RNA sequencing can reveal pathogenic mechanisms invisible to exome or genome analysis, including cryptic splicing, regulatory variants, and transcripts that escape nonsense-mediated decay. The findings establish RNA-seq as a decisive bridge between genotype and phenotype, uncovering functional consequences of genetic variation and redefining molecular diagnostics for mitochondrial and other neuro-metabolic diseases.

## Introduction

Mitochondrial dysfunction, predominantly characterized by defects in oxidative phosphorylation and impaired energy metabolism, gives rise to a spectrum of clinically diverse disorders collectively known as mitochondrial diseases. As a group, mitochondrial diseases constitute the most common class of inherited metabolic disorders, with an estimated incidence of approximately 1 in 5,000 individuals. Pathogenic variants in more than 400 genes in both nuclear and mitochondrial DNA have been implicated, giving rise to marked clinical and genetic heterogeneity. This variability, combined with extensive phenotypic overlap with other Mendelian disorders, renders molecular diagnosis particularly challenging.

The routine clinical implementation of whole exome sequencing (WES) has revolutionized the genetic diagnosis of mitochondrial diseases and facilitated the discovery of more than 200 novel disease genes^[1]^. Nevertheless, despite comprehensive genomic analyses, the diagnostic yield in suspected cases rarely exceeds 50%, leaving a substantial proportion of patients without a molecular diagnosis^[2]^. WES, by design, captures only exonic regions and therefore misses non-coding variants, structural rearrangements, and repeat expansions. While whole-genome sequencing (WGS), especially in its long-read form, can detect these variant types, its incremental diagnostic yield remains modest^[3]^. Assuming that most mitochondrial disorders are genetically caused, it is considered that genome-wide sequencing is able to detect the causal variant, but shifts the main bottleneck from variant detection to clinical prioritization and interpretation, emphasizing the need for integrating additional functional evidence into the diagnostic workflow. The discovery of millions of variants in the human genome resulted in an explosion of in silico prediction tools interpreting protein-coding variants^[4]^ and splice site variants^[5]^ with increasing accuracy; however, already the prediction of functional effects of near splice site variants becomes challenging, and the accuracy in predicting the consequences of deep intronic variants is still very low.

Current research indicates that approximately 30% of pathogenic variants are located in non-coding regions of the genome^[6, 7]^. Variants in the coding region may also affect gene expression or splicing. Multiplex splicing assays have revealed that around 10% of pathogenic exonic variants disrupt splicing^[8, 9]^, another category of alterations that remains challenging to predict accurately. Moreover, most predicted protein-truncating variants (PTVs) induce nonsense-mediated mRNA decay (NMD). NMD has a judgmental impact on the functional consequence. A large number of truncated proteins would remain functional, but NMD causes a loss-of-function. Roughly 30% of PTVs escape predicted NMD with a fundamental consequence on the clinical interpretation^[10–13]^. Consequently, without experimental evidence from patient biopsy material, many rare variants remain variants of uncertain significance (VUS) ^[14]^. This underscores the critical need and potential power of functional studies to elucidate the pathogenic potential of candidate variants.

RNA sequencing (RNA-seq) enables a comprehensive analysis of transcript abundance, facilitating the systematic detection of RNA phenotypes and providing allelic information across the entire transcriptome. These phenotypes include aberrant gene expression, abnormal splicing patterns, and monoallelic expression of rare variants. Identifying such anomalies can provide functional evidence for VUS in both coding and non-coding regions, facilitating their correct classification. Moreover, the analysis of RNA-seq data may also discover aberrant RNA phenotypes in genes not prioritized in the genomic evaluation. To date, RNA-seq has been applied to various patient cohorts and improved the diagnostic yield on average by 15%^[14]^.

Here, we report the implementation of RNA-seq within a national diagnostic program for mitochondrial disease at Beijing Children’s Hospital. We performed RNA-seq on fibroblasts from 140 paediatric patients who remained undiagnosed after WES and achieved a molecular diagnosis in 25% of cases, demonstrating that functional transcriptomics is a powerful addition to the diagnostic workflow. Our findings further highlight the limitations of current prediction tools for NMD and splicing and emphasize the necessity of empirical transcriptomic evaluation in the clinical interpretation of rare variants.

## Materials and Methods

### Study cohort

Patients were recruited between November 2017 and August 2023 from the Department of Neurology, Beijing Children’s Hospital, Capital Medical University, National Center for Children’s Health (China) through the Chinese Mitochondrial Disease Network (mitoC-Network; ChiCTR1900028101). Inclusion required a Mitochondrial Disease Criteria (MDC) score ≥2, or MDC <2 with a candidate mitochondrial gene^[15, 16]^, and no molecular diagnosis after whole-exome sequencing (WES). Skin biopsies were obtained from 140 paediatric patients (median age at biopsy 3.4 years; IQR 1.7–7.1; male:female ≈1.5:1). Written informed consent for genetic testing and research was obtained from legal guardians in accordance with the Declaration of Helsinki. The study was approved by the institutional ethics committee (2017-k-60).

Based on WES findings, patients were assigned to two groups: a candidate group (n=28), in which RNA-seq evaluated the pathogenicity of WES-identified variants of uncertain significance; and an unsolved group (n=112), in which RNA-seq was used to pinpoint candidate genes.

### WGS and WES reanalysis

Raw WES and WES/WGS data of each participant were reanalyzed. Reads were aligned to the human reference genome (GRCh38/hg38) using the Burrows-Wheeler Aligner (BWA 0.7.17-r1188) and MEM algorithm^[17]^. Variant calling for SNPs and indels was performed using the haplotyper algorithm of the Sentieon software suite (202010.02). ANNOVAR was used for variant annotation, including population frequency (1000 Genome Project^[18]^, ExAC^[19]^, gnomAD^[20]^, and the Beijing Children’s Hospital in-house population database), in silico pathogenicity predictions for missense variants (SIFT^[21]^, PolyPhen2^[22]^, LRT^[23]^, CADD^[24]^, REVEL^[25]^) and splice site variants (spliceAI^[26]^, MaxEntScan^[27]^, NNSplice^[28]^, and dbscSNV^[29]^). For the SpliceAI prediction, we used the conservative thresholds to ensure the specificity. A Δ score ≤ 0.01 was considered benign, and a Δ score ≥ 0.8 was considered pathogenic effects on splicing. Variants were classified according to the guidelines of the American College of Medical Genetics and Genomics and the Association of Molecular Pathology (ACMG/AMP)^[30]^. For structural variant detection, ExomeDepth(1.1.16)^[31]^ and the CNV kit^[32]^ were used for WES and WGS, respectively. Manta (version 1.5.0)^[33]^ and DELLY (V0.8.1)^[34]^ were applied to detect structural variations in WGS data. Structural variants were annotated using AnnotSV (v3.3).^[35]^

### Cell culture

Primary fibroblast cell lines obtained from patient skin biopsies (diameter 2–3 mm) were cultured in 1 mL culture medium with high-glucose DMEM (Life Technologies) supplemented with 20% FBS and 1% penicillin/streptomycin at 37 °C and 5% CO_2_. Culture media were changed every 2 days, and the skin flap was passaged approximately 10 days after implantation once confluence was reached.

### RNA isolation, library preparation, and sequencing

Fibroblast cells (>1*10^6^) were harvested in 1 mL TRIzol® reagent (Thermo Fisher Scientific). Total RNA was extracted using an AllPrep RNA Kit (Qiagen). RNA quality and integrity were assessed using an Agilent 2100 BioAnalyzer. PolyA-tailed mRNA was enriched with oligo (dT) magnetic beads, fragmented, and reverse-transcribed. Purified cDNA was subsequently subjected to end repair, A-tailing, and adapter ligation. 350–450-bp cDNA libraries were selected, PCR-amplified, and purified. High-quality libraries were sequenced as 150-bp paired-end runs on an Illumina NovaSeq 6000 platform (Illumina) to generate more than 40 million reads (12 GB raw data).

### RNA-seq alignment and variant calling

Low-quality bases and sequencing adapters were removed, and reads were aligned to the human GRCh38/hg38 genome assembly using STAR 2.7.6a^[36]^ with the twopassMode set to ”Basic” to enable sensitive junction discovery. Following the removal of duplicate reads, base quality score recalibration, and read splitting at the junction, variants from RNA-seq data were called using the Sentieon software suite (202010.02) Haplotyper algorithm with default parameters.

### Detection of aberrant RNA phenotypes

STAR-aligned BAM files were used to calculate gene-level counts based on NCBI Refseq (hg38.refseq.gff: GRCh38_20200919), which consisted of 39,225 genes by HTseq-count^[37, 38]^. Genes with 95th percentile fragments per kilobase of transcript per million mapped reads (FPKM) less than 1 were considered insufficiently expressed and were excluded from further analysis.

Gene count matrices from two batches (88 samples and 56 samples) of cultured skin fibroblast cells and related data from Genotype-Tissue Expression (GTEx) (cultured fibroblasts, skin not sun-exposed (suprapubic), skin sun-exposed (lower leg), and whole blood) were normalized and transformed for variance stabilization.

Aberrant expression (AE), aberrant splicing (AS), and monoallelic expression (MAE) were analyzed using the core module of DROP^[37]^. OUTRIDER(1.12.0)^[39]^ was used to identify aberrant candidate gene expression events using *p*-values (<0.005) and | Z-scores | (>3). All disease-related candidate events were subsequently validated using Integrative Genome Viewer^[40]^.

FRASER (1.7.0) was used to detect splicing outliers, which is based on count distribution and multiple testing corrections to reduce the number of calls by two orders of magnitude over the commonly applied Z-score cutoffs ^[37, 41]^. Exon-exon and exon-intron junctions with more than 10 reads in at least one sample were analyzed. Alternative acceptors (ψ5), alternative donors (ψ3), as well as splicing efficiencies at donors (θ5) and acceptors (θ3), were quantified to identify both alternative splicing and intron retention events. Outlier junctions were defined as those in splicing outlier genes, with p-values <0.05 and |Z scores| >2 and count ≥5.

For the detection of monoallelic expression, allelic RNA counts were extracted from the gVCF file based on the corresponding genomic variants identified from the WES data. Variants with a total depth of less than 10 reads in RNA-seq, fewer than five reads in WES, and variants with an allelic frequency of less than 0.3 in WES were filtered out. Monoallelic expression was determined using a negative binomial test applied to read counts using the tMAE package with the DESeq4MAE function^[37, 42, 43]^. Benjamini-Hochberg adjusted *p*-values <0.05 indicated significant MAE.

### WES and RNA integration

Sample concordance between RNA-seq and DNA was verified using Plink, with variants covered by >20 reads in > 90% of the samples^[44]^. Identity-by-descent (IBD) analysis was conducted to quantify the proportion of shared alleles (PI_HAT), with a threshold of PI_HAT > 0.8 threshold to confirm matches. For each individual, the RNA expression and splicing outliers were integrated using rare genomic variants (gnomAD minor allele frequency [MAF] < 0.01). To refine the event prioritization in our RNA-seq diagnostics pipeline, inheritance-specific filters for splicing outliers were integrated. Candidate splice anomalies were filtered as follows: for dominant disease-associated genes: (1) p-value < 0.01, |deltaPsi| > 0.1, and gene with potentially splice-disrupting variants (minimum distance ≤200 between variant and splice junction, or located within splice regions); (2) p-value < 0.01, expression Z-score < -2, and gene with potentially splice-disrupting variants; (3) p-value < 0.005, |deltaPsi| > 0.1. For recessive disease-associated genes, (1) *p*-value < 0.005, |deltaPsi| > 0.5; (2) *p*-value<0.01, |deltaPsi| > 0.1, and genes with at least one variant were classified as pathogenic/likely pathogenic/uncertain.

### Systematic literature review

We searched PubMed, ISI Web of Science, and Scopus (January 2017–September 2024) using: (“Rare disease” OR “Mendelian disease” OR “Mendelian disorders”) AND (“transcriptome” OR “transcriptome sequencing” OR “RNA-seq” OR “RNA sequencing”). Inclusion criteria were original cohort studies applying RNA-seq for Mendelian disease diagnostics. Reviews, meta-analyses, case reports, and unrelated articles were excluded. Data were extracted on cohort size, tissue, analytical pipeline, and diagnostic yield. The analysis was patient-based.

### Data availability statement

The data needed to reproduce the figures are available upon request. Anonymized data not provided in the article may be shared at the request of any qualified investigator to replicate the procedures and results.

## Results

### Study design and patient characteristics

Within the last five years, the Chinese Mitochondrial Disease Network (mitoC-Network) recruited 1,405 patients suspected of having a mitochondrial disease and performed genome-wide molecular screening to identify disease-causing variants. This was achieved in approximately 45% of cases, and hence no genetic diagnosis could be made for the majority of patients. To support routine diagnostic testing, we established fibroblast cell lines from 140 genetically unsolved patients for functional studies. The most frequent clinical presentation in this cohort of patients was Leigh syndrome (36%), followed by non-specific phenotypes (29%) and movement disorders (9%) (Fig. 1A).

**Fig. 1:**
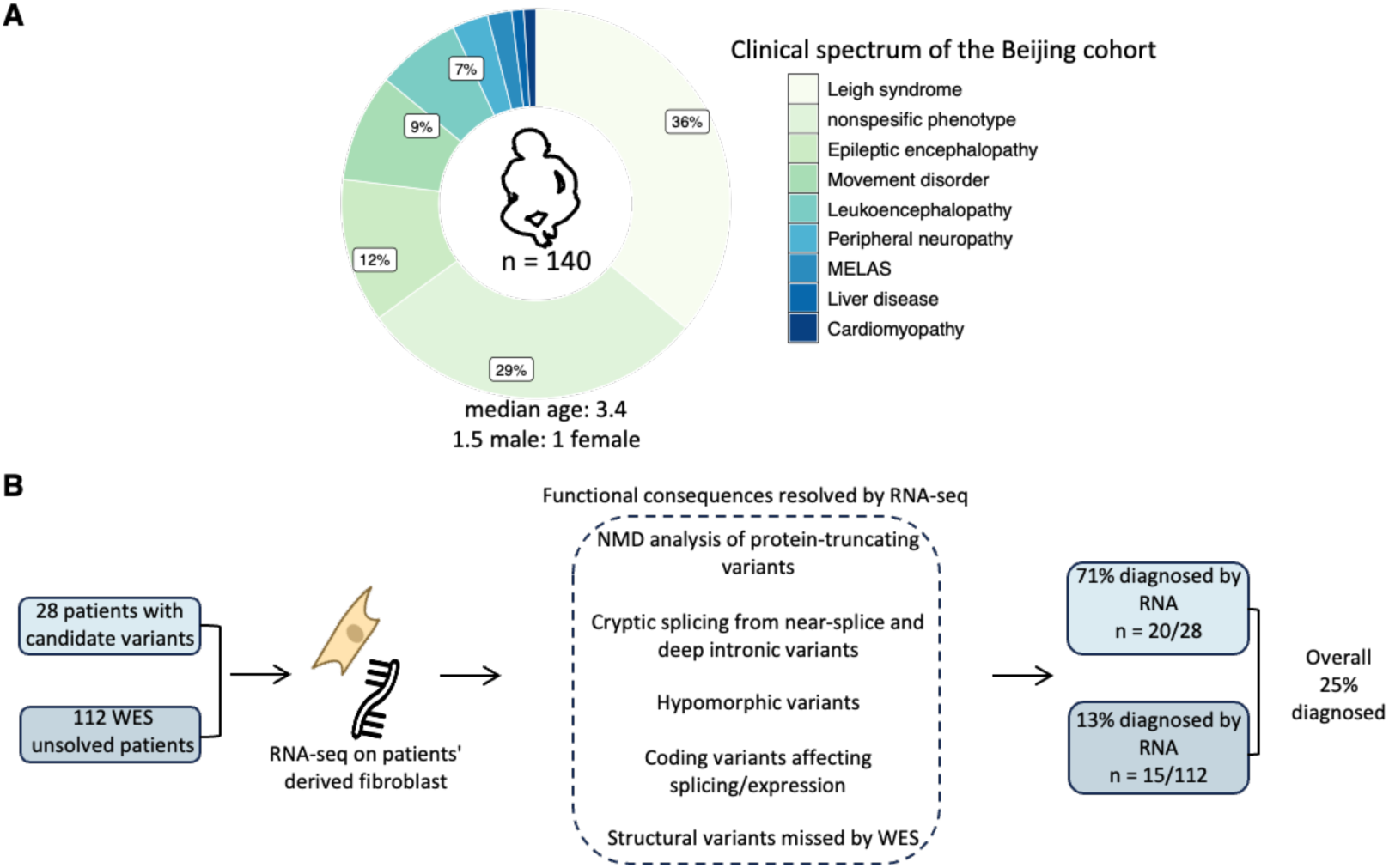
RNA-based diagnostic workflow for WES inconclusive pediatric patients with suspected mitochondrial disorders. (A) Clinical characteristics of 140 pediatric patients in the cohort. (B) Study design and outcome. Patients were categorized into two groups: candidate (n=28), in whom WES had prioritized variants and RNA-seq was used to assess the functional effect (molecular diagnosis in 20/28, 71.4%); and unsolved (n=112), with no prioritized variants, in whom RNA-seq was used to identify the candidate genes with aberrant RNA phenotypes followed by WES reanalysis or WGS (diagnosis in 15/112, 13.4%). The overall diagnostic yield of 25% (35/140) was achieved. WES: whole-exome sequencing, NMD: nonsense-mediated mRNA decay.

RNA-seq was conducted on a per-individual basis. Each sample was subjected to a single assay at a median sequencing depth of 94.5 million reads, ranging from 74.5 to 133.4 million reads, using a non-strand-specific protocol that utilized automated protocols to minimize sample handling and ensure high reproducibility of results (see Methods). After alignment, RNA-seq data were analyzed using the computational workflow DROP to search for aberrant expression, aberrant splicing, or monoallelic expression of a rare variant. RNA-seq results were then integrated with genomic and clinical data to guide variant interpretation (Fig. 1B).

### Evaluation of variants of uncertain significance

Among the 140 patients, 28 harboured candidate variants of uncertain significance in genes consistent with their clinical presentation. These included near splice-site (n = 10, +/- 3 to 153) and protein-truncating variants (PTVs; n = 15) (Fig. 2A; Supplementary Table 1).

**Fig. 2:**
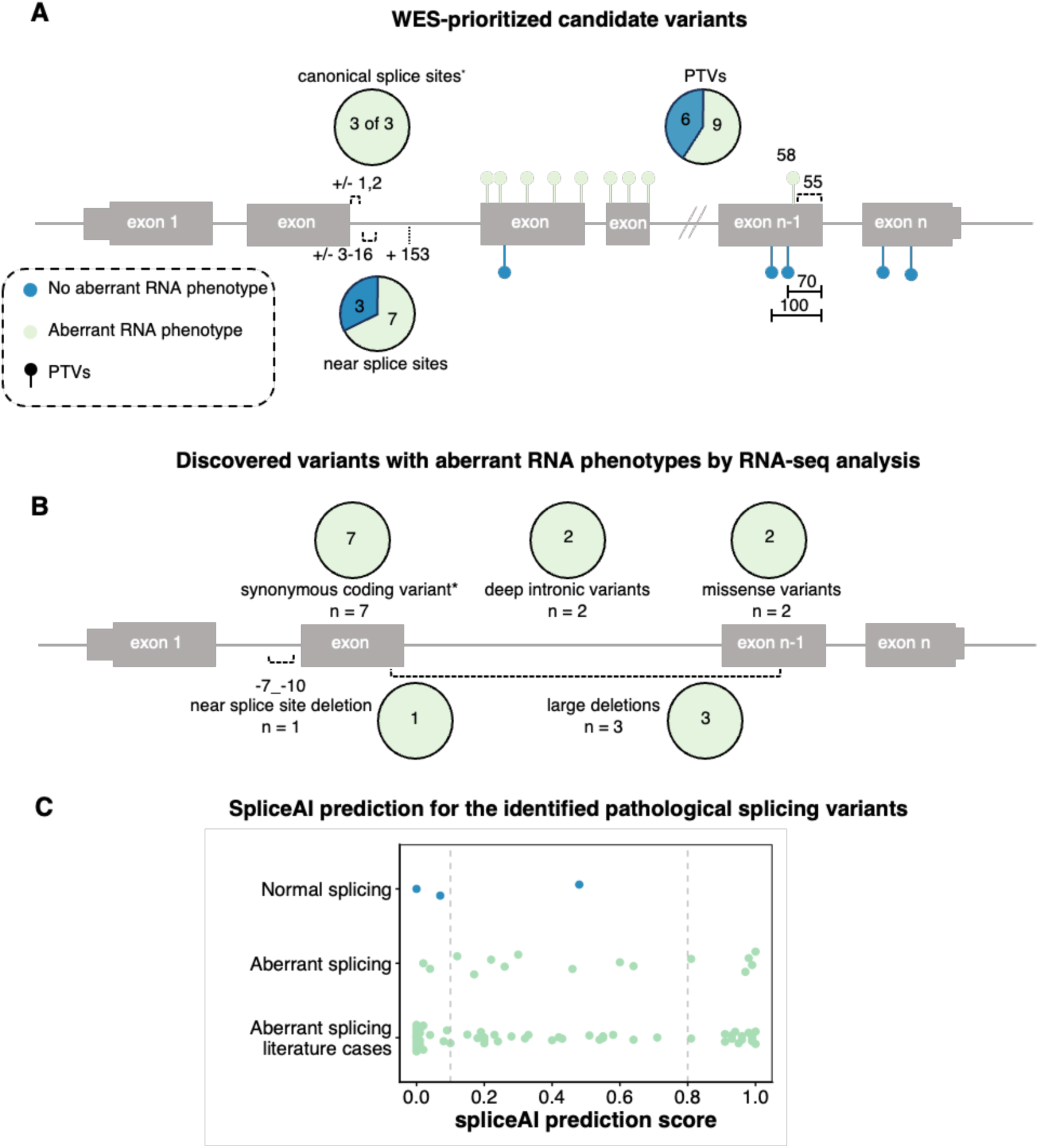
Variant types causing aberrant RNA phenotypes and the added value of RNA-seq over prediction tools. (A) The RNA phenotypes in association with the WES-prioritized candidate variants are shown. The dataset comprises 30 variants across 28 patients. The number in the pie chart shows the number of cases. With the exception of the canonical splice site variants (2 variants observed in 3 cases), the count of the cases equals the count of the variants. The numbers on the gene structure represent the nucleotide distance of each variant to the nearest exon junction. One missense variant (no RNA phenotype), one in-frame deletion (no RNA phenotype), and one large deletion (aberrant expression) were not shown in the graph, given the presence of only one variant per category. (B) Shown are the variant types identified in 15 cases that were genetically solved after RNA-seq. The dataset includes 9 variants across 15 patients. Numbers indicate cases. For all classes except the synonymous variant (1 variant observed in 7 cases), the number of variants equals the number of cases. (C) SpliceAI prediction for variants with and without aberrant RNA splicing (based on RNA-seq). The dataset includes 18 variants from panels A and B: 11 near splice site variants (10 from panel A and 1 from panel B), 2 canonical splice, 1 synonymous variant, 2 missense variants, and 2 deep intronic variants, and 64 variants causing aberrant splicing reported in literature. PTVs: protein truncating variants, WES: whole-exome sequencing.

RNA-seq provided functional validation for 20 of these 28 cases. Among the 15 predicted PTVs, NMD was confirmed in nine, while six showed no evidence of NMD. In one case, the affected gene (*PIGM*) contained only one exon. One affected gene (*SCN1A*) was not expressed in fibroblasts. In another case, premature termination codons (PTCs) occurred within the final 55 nucleotides of the penultimate exon of the gene (*COA6*); it was not predicted to cause NMD. However, in two cases with variants located 70 and 100 nucleotides upstream of the last splice junction in the penultimate exon of the genes (*IBA57*, *GLRX5*), NMD was predicted but not observed. In other cases, involving variants in the same region, we observed NMD. This highlights that predicting NMD in this region is inaccurate and that RNA-seq is needed to determine the functional consequences of such variants. In one patient, a frameshift variant occurs in the second of nine exons in the main *NDUFAF6* transcript, which should result in NMD. However, RNA-seq revealed that *NDUFAF6* expression levels were within the normal physiological range. Manual inspection revealed a high number of well-expressed *NDUFAF6* transcript isoforms and showed that the variant is outside the coding region in the 5’ UTR in 90% of them. It is not possible to conclude from the short-read data whether there is NMD of the isoform in which the variant is located within the coding region. Despite this, by leveraging established fibroblasts, studies of respiratory chain activities provided functional evidence for the pathogenicity of compound heterozygous PTVs in three patients (Supplementary Table 1). In conclusion, RNA-seq provided a loss-of-function characterization for the nine PTVs, thereby confirming their pathogenicity and aiding in diagnosis.

In 13 cases, the genome analysis identified variants that may affect splicing: three canonical splice site variants, and nine near splice site variants in positions +/–3–16, plus one variant at position +153. RNA-seq analysis confirmed a clear splice defect in 10 of 13 cases. Importantly, five of seven near-splice-site variants were misclassified by SpliceAI, demonstrating the diagnostic value of empirical RNA-seq validation (Supplementary Table 1).

In contrast, an in-frame deletion in *HSD17B4* and a missense variant in *POLG* showed no RNA abnormalities, while a large deletion in another gene was detected through reduced expression. In summary, RNA-seq analysis helped to establish a definitive molecular diagnosis in 20 out of 28 candidate cases, corresponding to a diagnostic rate of 71%.

### Discovery of disease-causing variants by RNA-seq

In the second group of 112 unsolved cases for which no candidate genes could be identified, integrating genomic data with aberrant RNA phenotypes and their clinical interpretation resulted in a molecular diagnosis for 15 patients, which we will discuss in detail below. The variants causing aberrant RNA phenotypes included: a common synonymous variant causing a splice defect (in seven cases); three intragenic deletions that were either not covered or called by WES; two predicted missense variants causing aberrant splicing or expression; two deep intronic variants and a deletion of -7 to -10 near splice sites that were missed by WES (see Supplementary Table 1, Fig. 2B). SpliceAI failed to predict most of the splice defects, underscoring the empirical power of RNA-seq in variant interpretation (Fig. 2C).

### A synonymous *ECHS1* variant causes aberrant splicing in seven patients with Leigh syndrome

RNA-seq detected underexpression of *ECHS1* in seven unrelated patients with Leigh syndrome (Fig. 3), all heterozygous for the synonymous variant c.489G>A (p.Pro163=) in trans with known pathogenic variants (p.Met103Thr, p.Thr266Ile, p.Thr266Ala, p.Glu274Lys, p.Ala278Thr, and p.Phe279Leu). The variant, located 25 bp from the donor and 74 bp from the acceptor splice site, was not predicted to affect splicing and expression. However, we detected partial skipping of the exon containing the variant (supported by 4% of reads), leading to a frameshift (*p.Phe139ValfsTer65*). This is predicted to cause NMD, which explains the under-expression of *ECHS1*. Despite NMD, exon 4 skipping was supported by 70 reads across all *ECHS1* patients. Overall, *ECHS1* expression was reduced by approximately 20% across patients, indicating that ∼40% of transcripts derived from the allele carrying the synonymous variant underwent NMD. This, together with the splice defect, leaves only half of the transcribed synonymous allele functional. This observation is further supported by monoallelic expression of the missense variants in trans in all seven cases. The observed ratio of alleles with 1/3 of the synonymous variant and 2/3 of the missense variants perfectly fits with the observed NMD quantification. Altogether, these data show that 25% of intact *ECHS1* is not enough for preventing the manifestation of Leigh syndrome, while 50% of *ECHS1* expression level is tolerated as observed in homozygous carriers of the synonymous variant. The c.489G>A variant has a high allele frequency of 1.05% in the East Asian population (Fig. 3C), suggesting a regional founder effect.

**Fig. 3.**
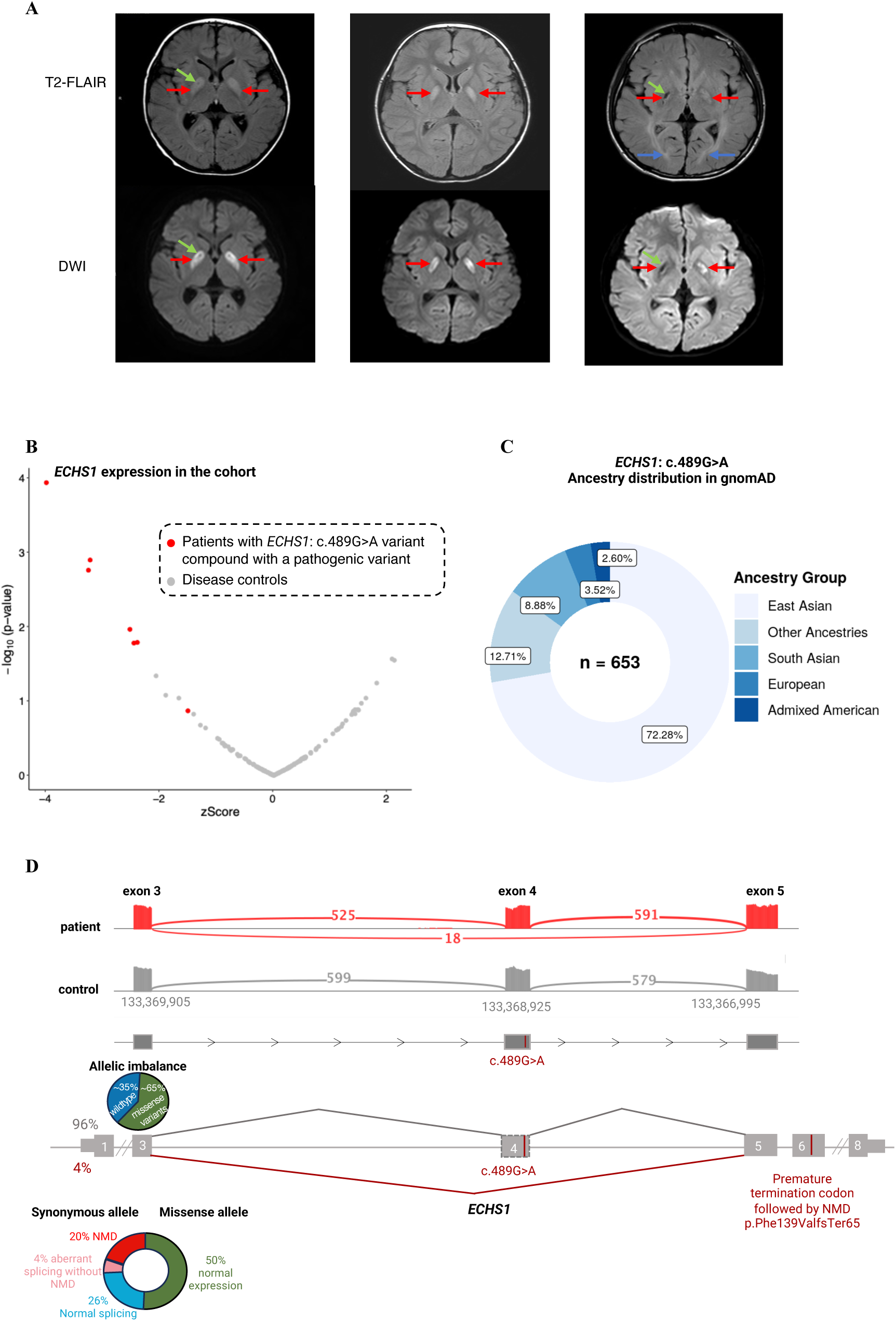
Aberrant expression of *ECHS1* in seven unsolved Leigh syndrome cases harbouring the common synonymous variant c.489G>A (p.Pro163=). (A) MRI images of three of the seven patients. Arrows indicate the positions of abnormal signals. Bilateral globus pallidus (red arrows) involvement was a common feature; case 2 showed punctate necrosis (green arrows), case 5 showed patchy necrosis (green arrows), and necrotic lesions were not accompanied by diffusion restriction (DWI). In addition to globus pallidus involvement, the bilateral optic radiation was also affected in case 5 (blue arrows). (B) Volcano plot of *ECHS1* gene expression across all the cases. Seven *ECHS1* diagnosed cases carrying c.489G>A variant in trans with other pathogenic variants are labeled in red. (C) Ancestry distribution of *ECHS1:* c.489G>A variant based on the gnomAD database shows the high frequency of the variant in the East Asian population. (D) *ECHS1* sashimi plots of fibroblast-derived RNA-seq data from an *ECHS1* patient, and one control, in red and grey respectively. The numbers on the exon-connecting lines show the number of unnormalized reads spanning the exons. Partial skipping of exon 4 (in 4% of the reads) is observed in the patient. The position of the c.489G>A variant in exon 4 is indicated. The schematic presentation of aberrant (red) and normal (grey) splicing is depicted at the bottom. The skipping of exon 4 due to c.489G>A variant results in a frameshift and premature termination codon (p.Phe139ValfsTer65) in exon 6. The average allelic imbalance across all seven missense variants present in trans with the synonymous variant is shown in a pie chart. The relative allelic expression of missense and synonymous variants in *ECHS1* transcripts is depicted in a donut chart based on the mean value across all patients. gnomAD: genome aggregation database.

*ECHS1* encodes short-chain enoyl-CoA hydratase 1, which is involved in mitochondrial valine metabolism and short-chain fatty acid beta-oxidation. The clinical presentation of *ECHS1* deficiency (OMIM #616277) is heterogeneous^[45, 46]^, but often includes severely delayed psychomotor development, neurodegeneration, increased lactic acid levels, and brain lesions in the basal ganglia consistent with Leigh syndrome. The clinical findings in these seven cases were consistent with a diagnosis of *ECHS1* deficiency. All individuals presented with similar clinical manifestations and neuroimaging features characteristic of Leigh syndrome (Fig. 3A), alongside relatively stable conditions. None had echocardiographic signs of cardiomyopathy. Metabolic testing performed on four patients revealed elevated urine concentrations of 2,3-dihydroxy-2-methylbutyric acid in all of them and an elevated concentration of S-(2-carboxypropyl) cysteamine in three, further supporting the diagnosis (see Supplementary Table 2). Treatment was modified accordingly with a valine-restricted diet and acetylcysteine supplementation, which were initiated after diagnosis^[47, 48]^.

### RNA-seq-guided detection of cryptic deletions in *PANK2* and *SERAC1*

A patient exhibiting neurodevelopmental delay and regression, as well as dystonia, underwent a brain magnetic resonance imaging (MRI) scan, which revealed bilateral abnormalities in the globus pallidus, indicating the ’eye of the tiger’ sign (Fig. 4B). This finding supported the diagnosis of neurodegeneration with brain iron accumulation (NBIA). Trio-WES identified a rare maternal heterozygous missense variant (p.Arg440Pro) in *PANK2* (Fig. 4A), which has been previously reported in association with NBIA ^[49, 50]^. RNA-seq revealed significantly reduced *PANK2* expression levels (Fig. 4C). The maternal variant was monoallelically expressed, indicating the loss of expression of the paternal allele (Fig. 4D). Trio-WGS identified a paternal intragenic deletion involving the terminal exon, consistent with compound heterozygosity and confirming *PANK2*-related NBIA (OMIM #234200, Fig. 4A-D).

**Fig. 4.**
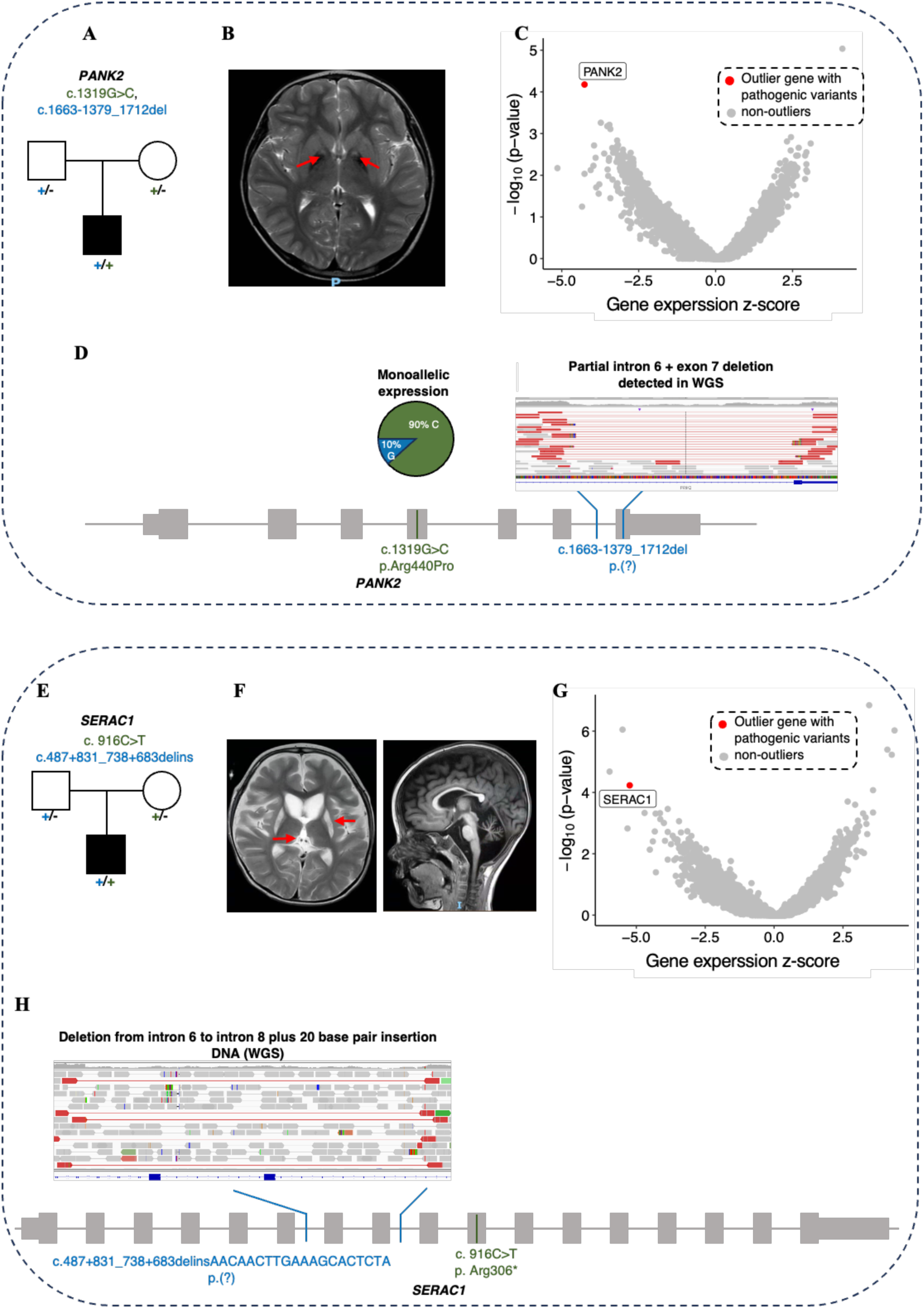
Aberrant expression caused by large intragenic deletions, undetected by WES. (A-D) *PANK2* patient; (E-H) *SERAC1* patient. (A) Family pedigree; *PANK2* compound heterozygous pathogenic variants identified after RNA-seq and trio-WGS are indicated. (B) Brain MRI in the T2-weighted image. The red arrow indicates bilateral abnormalities in globus pallidus, also known as “eye of the tiger” sign. (C) Gene-level volcano plot, showing the significance versus z-score of the gene expression in the patient, with the *PANK2* highlighted as the most significant underexpression outlier. (D) Schematic representation of the *PANK2* gene structure, showing the position of the missense variant (chr20:3912541G>C, c.1319G>C) and large deletion (chr20:3921864-3923293del) identified in the patient. The monoallelic expression of the missense variant (c.1319G>C) in RNA-seq data (RNA: alt total=85/94 (90%)) indicates the reduced expression of the allele carrying the large deletion. IGV from the mapped reads of the patient’s WGS data shows the heterozygous intragenic deletion (chr20:3921864-3923293del). (E) Family pedigree; *SERAC1* compound heterozygous pathogenic variants identified after RNA-seq and trio-WGS are indicated. (F) Brain MRI in T2-weighted images showed bilateral basal ganglia and thalamus abnormalities (red arrows), and cerebral and cerebellar atrophy. (G) Gene-level volcano plot, showing the significance versus z-score of the gene expression in the patient. *SERAC1 i*s highlighted as the underexpressed outlier harboring two pathogenic variants. (H) Schematic representation of the *SERAC1* gene structure, showing the position of the large deletion-insertion (chr6:158142373-158145953delinsTAGAGTGCTTTCAAGTTGTTTG, c.487+831_738+683delinsAACAACTTGAAAGCACTCTA) and stop-gain variant (chr6:158128207 G>A, c. 916C>T) identified in the patient. IGV from the mapped reads of the patient’s WGS data shows the heterozygous chr6:158142373-158145953delinsTAGAGTGCTTTCAAGTTGTTTG. *SERAC1* is transcribed from the reverse strand but, for consistency, is depicted in the forward orientation in the schematic. WES, whole-exome sequencing; WGS, whole-genome sequencing; IGV, Integrative Genomics Viewer.

A boy had presented with liver dysfunction, failure to thrive, neurodevelopmental delay, and hearing impairment in infancy. Later, epilepsy developed with a first status epilepticus, and since then, developmental regression has been observed. Metabolic evaluation revealed lactic acidosis in the blood and elevated urinary 3-methylglutaconic acid excretion. On brain MRI, abnormalities in the basal ganglia and thalamus were detected, which were interpreted as Leigh syndrome, together with generalized cerebral and cerebellar atrophy (Fig. 4F). His sibling had died from lactic acidosis. Initial WES identified heterozygosity for a predicted pathogenic PTV (p.Arg306*) in *SERAC1* (Fig. 4E). RNA-seq analysis revealed strongly reduced *SERAC1* expression levels (Fig. 4G), less than expected from monoallelic NMD. Consequently, trio-WGS revealed a large deletion-insertion structural variant spanning from intron 6 to intron 8 in the paternal genome (Fig. 4H). The SERAC1 protein is involved in the remodelling of phosphatidylglycerol 36:1 with the effect of abnormal cardiolipin species in mitochondrial membranes^[51]^. Pathogenic variants within *SERAC1* are associated with 3-methylglutaconic aciduria, hepatopathy, deafness, encephalopathy, and Leigh-like syndrome (MEGHDEL, OMIM #614739), which is consistent with the patient’s clinical presentation and in line with the genetic diagnosis.

### Pseudoexon creation by deep intronic variants in *WARS2* and *SUCLG1*

A boy presented with nystagmus, ataxia, seizures, developmental regression, and elevated blood lactate. A brain MRI revealed mild cerebral atrophy (Fig. 5B). Trio-WES did not identify any candidate gene. RNA-seq revealed aberrant splicing that created a pseudoexon in intron 1 of *WARS2* by activating a cryptic splice site (Fig. 5C). RNA-seq variant calling detected a homozygous deep intronic variant (c.90+268C>T) located within the pseudoexon. Trio-WGS confirmed that this was the only rare variant in intron 1 and was inherited from both parents (Fig. 5A). It is predicted that the creation of the pseudoexon will lead to a stop-gain codon after eight amino acids and NMD. However, no NMD was detected. *WARS2* encodes mitochondrial tryptophanyl-tRNA synthase, which catalyzes the aminoacylation of mitochondrial tRNA with tryptophan. Biallelic mutations in *WARS2* are associated with neurodevelopmental disorders, abnormal movements, and lactic acidosis with or without seizures (NEMMLAS, OMIM #617710), symptoms in line with the patient’s presentation.

**Fig. 5.**
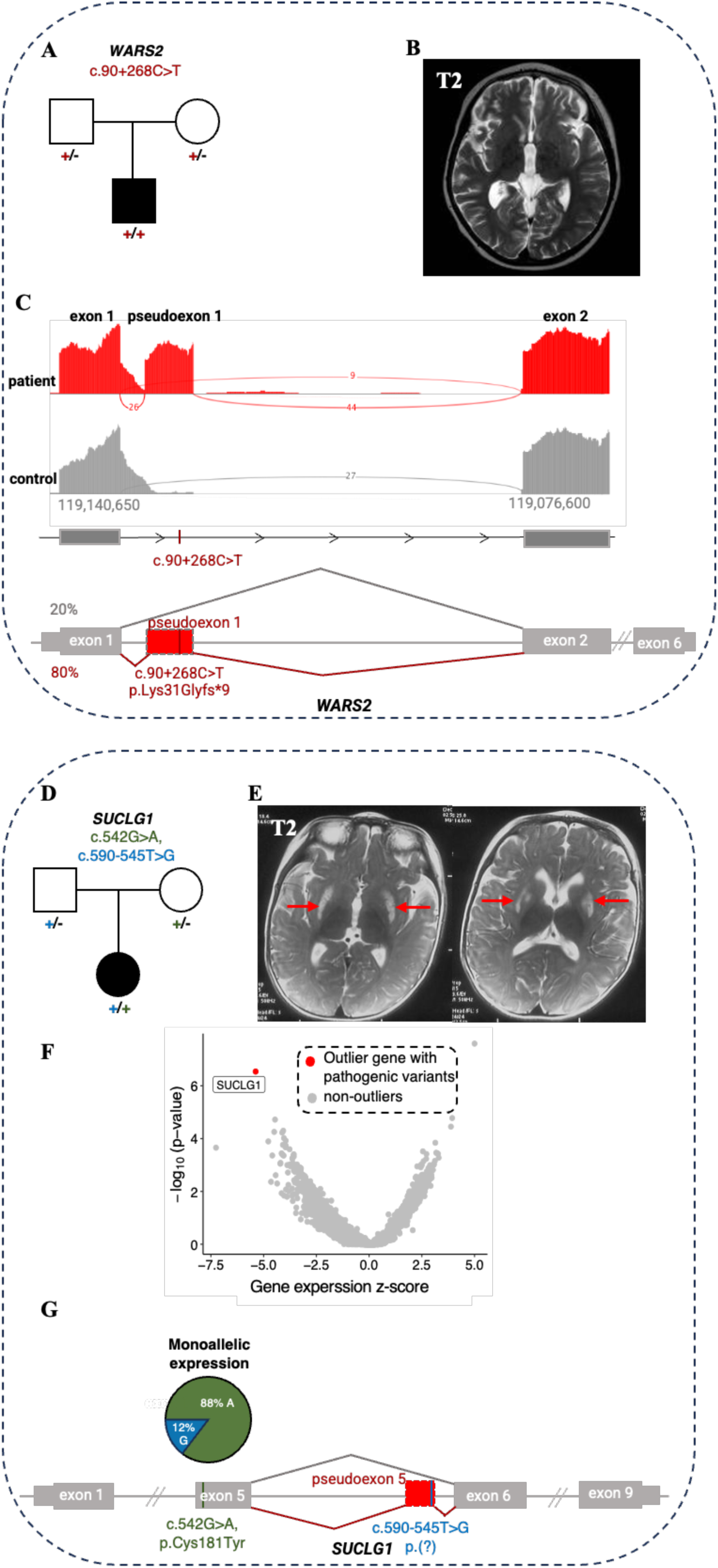
Aberrant splicing due to deep intronic variants, not coerced by WES data. (A-C) *WARS2* patient; (D-G) *SUCLG1* patient. (A) Family pedigree; *WARS2* homozygous deep intronic variant (chr1:119140287G>A, c.90+268C>T) identified after RNA-seq and trio-WGS is indicated. (B) Brain MRI in T2-weighted image, showing mild cerebral atrophy-like changes. (C) *WARS2* sashimi plots of fibroblast-derived RNA-seq data from the *WARS2* patient, and one control, in red and grey respectively. The numbers on the exon-connecting lines show the number of unnormalized reads spanning the exons. Creation of a pseudoexon in intron 1 due to a homozygous c.90+268C>T deep intronic variant is observed in the patient. The schematic presentation of aberrant (red; ∼80% of the reads) and normal (grey; ∼20% of the reads) splicing in the patient is depicted at the bottom. Inclusion of the pseudoexon introduces a frameshift with a premature termination codon within the pseudoexon (p.Lys31Glyfs*9). *WARS2* is transcribed from the reverse strand but, for consistency, is depicted in the forward orientation in the schematic. (D) Family pedigree; *SUCLG1* compound heterozygous variants identified after RNA-seq and trio-WGS are indicated. (E) Brain MRI in T2-weighted images, showing symmetrical basal ganglia lesions (red arrows) and cerebral atrophy. (F) Gene-level volcano plot, showing the significance versus z-score of the gene expression in the patient, with *SUCLG1* highlighted as the most significant underexpressed outlier. (G) Schematic representation of the *SUCLG1* gene structure, showing the missense variant (chr2:84441094C>T, c.542G>A) and deep intronic variant (chr2:84433980A>C, c.590-545T>G). The monoallelic expression of the missense variant (c.542G>A) in RNA-seq data (RNA: alt/total=88%) indicates the reduced expression of the allele carrying the intronic variant. Aberrant splicing in association with the deep intronic variant results in the creation of a pseudoexon in intron 5 of the gene (shown in red). *SUCLG1* is transcribed from the reverse strand but, for consistency, is depicted in the forward orientation. WES, whole-exome sequencing; WGS, whole-genome sequencing.

A girl presented with hypotonia, developmental delay and regression. She also had an elevated blood lactate concentration and showed mild but unequivocally increased urinary excretion of methylmalonic acid (MMA). Brain MRI showed symmetrical basal ganglia lesions and bilateral lateral ventricle broadening (Fig. 5E). The patient was clinically classified to have Leigh syndrome. RNA-seq analysis revealed a significant decrease in *SUCLG1* expression (Fig. 5F). A rare heterozygous predicted pathogenic missense variant (c.542G>A, p.Cys181Tyr) showed monoallelic expression (Fig. 5G). Furthermore, aberrant splicing of a pseudoexon in intron 5 was detected with very few reads, indicating NMD of this splice isoform. RNA-seq variant calling revealed a variant in the pseudoexon that likely activated the cryptic splice donor site (Fig. 5G). Trio-WGS confirmed that the deep intronic variant c.590-545T>G is *in trans* with the missense variant (Fig. 5D). Biallelic mutations in *SUCLG1*, encoding the alpha subunit of succinate–coenzyme A ligase, have been associated with Leigh syndrome and methylmalonic aciduria (OMIM #245400), a clinical presentation that is fully consistent with the symptoms of the patient.

### A homozygous missense variant in *CARS2* causes aberrant splicing

A boy presented with severe, progressive, drug-therapy-refractory epileptic seizures. He showed psychomotor developmental regression and was clinically diagnosed with developmental and epileptic encephalopathy. His siblings had exhibited similar symptoms and had died (Fig. 6A). Brain MRI of the index case revealed massive cerebral and cerebellar atrophy (Fig. 6B), and electroencephalography (EEG) showed significant epileptic activity. Trio-WES identified a homozygous missense VUS in *CARS2*. RNA-seq analysis revealed aberrant splicing of *CARS2*, resulting in less than 5% of the primary transcript being retained (Fig. 6C). The coding VUS located 10 base pairs downstream of the canonical exon 14 acceptor splice site significantly reduced its recognition, resulting in alternative usage of a weak acceptor in intron 13. This resulted in a final exon without further splicing, thereby omitting exons 14 and 15. This alternative exon usage is predicted to create a termination codon after the insertion of 25 amino acids (p.Tyr473GlnfsTer26). Despite the splice defect, overall gene expression was unaffected. Biallelic mutations in *CARS2*, which encodes the mitochondrial cysteinyl-tRNA synthetase, cause combined oxidative phosphorylation deficiency type 27 (OMIM #616672), characterized by developmental regression and epileptic encephalopathy, thus an excellent match to the symptoms of the patient.

**Fig. 6.**
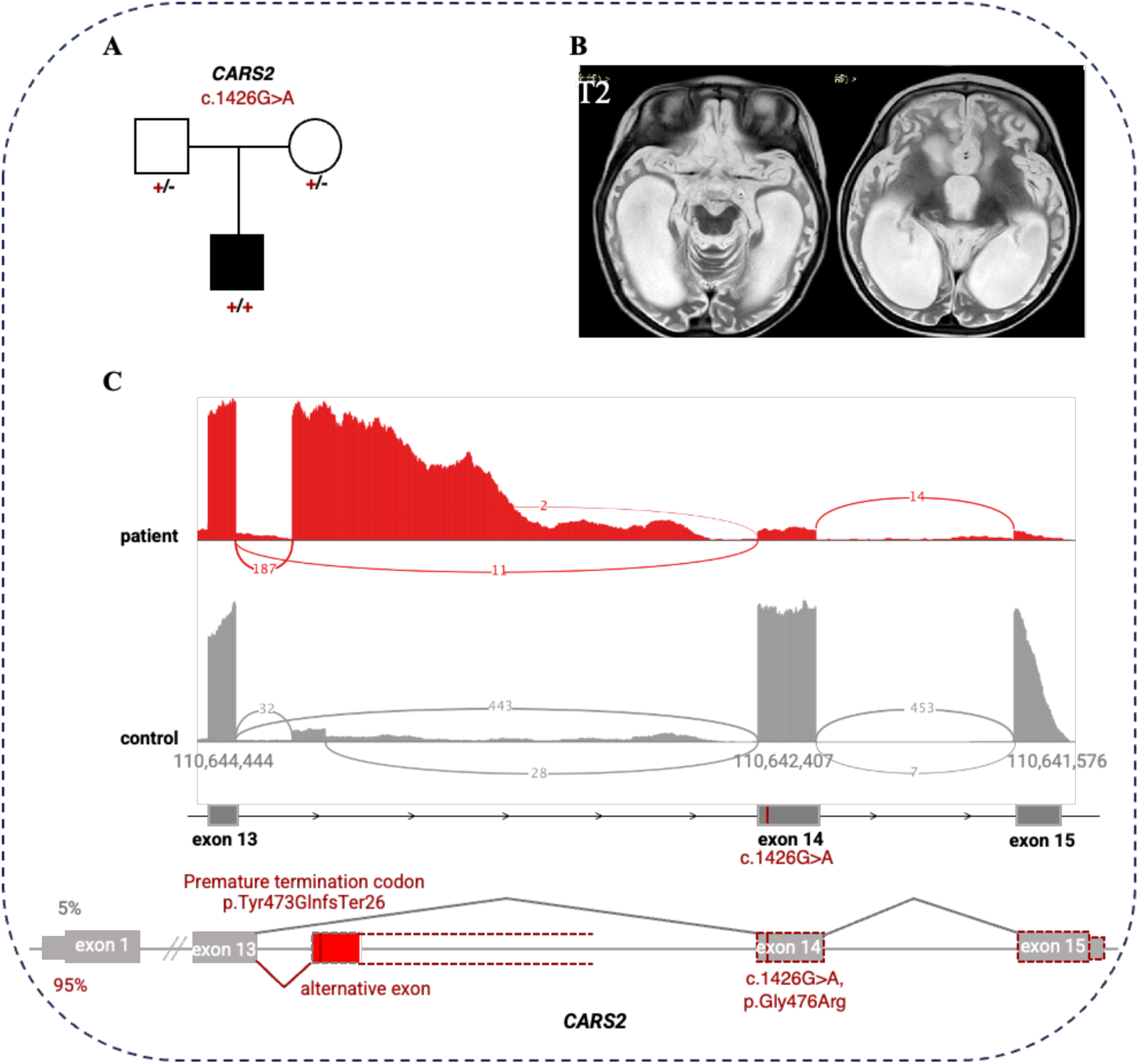
Homozygous missense variants in *CARS2* cause aberrant splicing. (A) Family pedigree; *CARS2* homozygous missene variant (chr13:110642512C>T, c.1426G>A) identified after RNA-seq and WES renalaysis is indicated. (B) Brain MRI shows severe cerebral and cerebellar atrophy. (C) *CARS2* sashimi plots of fibroblast-derived RNA-seq data from the *CARS2* patient, and one control, in red and grey respectively. The numbers on the exon-connecting lines show the number of unnormalized reads spanning the exons. Activation of the alternative splice site and the intron retention is observed in the patient. The schematic presentation of aberrant (red; ∼95% of the reads) and normal (grey; ∼5% of the reads) splicing in the patient is depicted at the bottom. The c.1426G>A variant in exon 14 reduced the usage of the exon 14 acceptor splice site and activated the alternative splice site within the intron, resulting in a premature termination codon and reduced expression of exons 14 and 15. The total expression of the gene is not affected. *CARS2* is transcribed from the reverse strand but, for consistency, is depicted in the forward orientation. WES, whole-exom sequencing.

In summary, the integration of RNA-seq data from fibroblasts with the genomic and clinical information of 140 patients suspected of having a mitochondrial disorder resulted in a molecular diagnosis in 35 cases, yielding an overall diagnostic rate of 25% (Fig. 1, Supplementary Table 1). With the exception of three cases, all of the identified causal genes were already associated with mitochondrial diseases (Supplementary Table 1).

### Comparison with published RNA-seq studies

Including this cohort, 16 studies have reported RNA-seq implementation for Mendelian disorders (Supplementary Table 3), encompassing 1,049 patients and 227 molecular diagnoses, for an average diagnostic yield of 22%^[14, 52–66]^. Diagnostic success varied by tissue type and disorder: highest in muscle (34–38%) in 138 patients with neuromuscular disorders^[53, 54, 63]^, intermediate in fibroblasts (10–33%) in 501 patients across six cohorts with diverse phenotypes, and lowest in blood (5–8%) in 178 patients with suspected rare diseases^[57, 66]^. One study performed RNA-seq analysis of induced neurons derived from skin fibroblasts and achieved a diagnostic efficiency of 25.4% (18/71) in patients with primary neurological phenotypes^[65]^.

Across 233 curated pathogenic variants with aberrant RNA phenotypes, half resided in non-coding regions and half in coding regions (Fig. 7, Supplement Table 4). The most common non-coding variants were deep intronic variants (19%), whereas missense (15%) and frameshift (12%) variants predominated among coding alterations

**Fig. 7.**
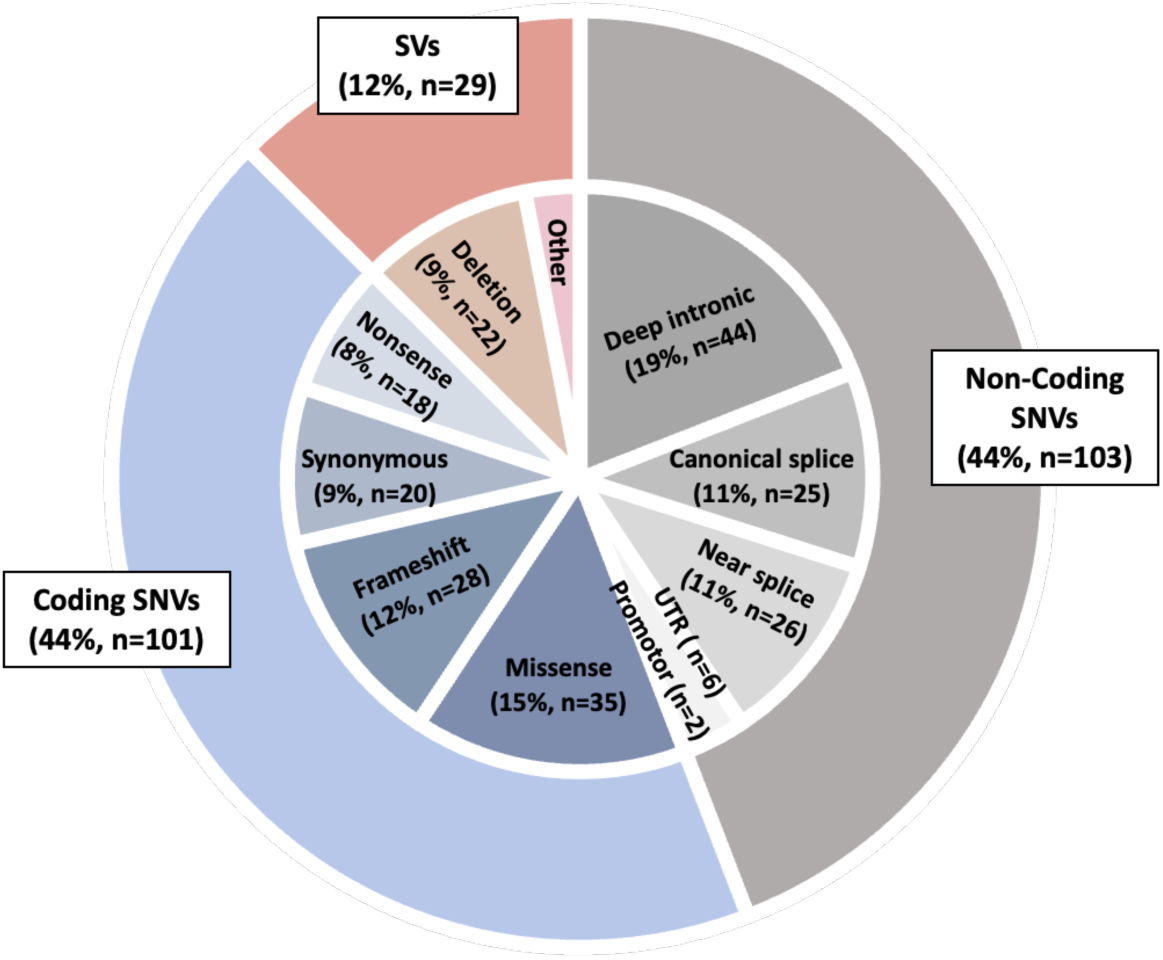
Variant types underlying the aberrant RNA phenotypes curated from this study and previous reports. The figure is patient based (the same variant in X patients counted X times). A total of 233 variants with aberrant RNA-seq events, including non-coding (n=103, 44%), coding single-nucleotide variants (SNVs) (n=101, 44%), and structural variants (SVs) (n=29, 12%), were identified. Non-coding SNVs included deep intronic variants (n=44), canonical splice sites (n=25), near splice sites (±3 to ±10 bp) (n=26), UTR (n=6), and enhancer (n=2). Coding SNVs included missense (n=35), frameshift (n=28), synonymous (n=20), and nonsense (n=18). Among the structural variants (SVs), deletion was the most common (n=22), followed by complex SV (n=2), inversion-deletion, inversion, duplication, insertion, and insertion-deletion.

## Discussion

This study demonstrates that integrating RNA sequencing into the genomic diagnostic workflow substantially enhances the resolution of genetic testing for neuro-metabolic diseases. Unlike the genome, the transcriptome is cell-type specific. Fibroblast cell lines, used in this study, offer an optimal balance between accessibility and the breadth of expressed genes^[14]^. In our fibroblast-derived transcriptome dataset, 15,536 genes were detected, covering 67% of Mendelian disease-associated genes and 90% of genes implicated in mitochondrial disorders. By combining RNA-seq with WES and WGS in a large, clinically well-characterized paediatric cohort, we achieved a molecular diagnosis in 25% of previously unresolved cases. Beyond identifying additional pathogenic variants, our data provide functional insights into how diverse classes of genomic variants—coding, non-coding, and structural—disrupt gene expression, splicing, or transcript stability.

### Functional validation of variants of uncertain significance

Genome-wide sequencing has transformed rare disease diagnostics but has also revealed the complexity of variant interpretation. The large number of variants identified in each patient demands rigorous classification frameworks such as ACMG/AMP, supported by curated resources including ClinGen, ClinVar, and disease-specific databases. Such strict standards minimize false-positive diagnoses but also increase the number of candidate variants of uncertain significance (VUS)^[67]^. In ClinVar, more than half of likely pathogenic variants are protein-truncating variants (PTVs), whereas missense variants account for approximately 25–30%^[68]^. This discrepancy reflects the greater predictability of loss-of-function variants compared with missense changes rather than their true frequency in the population. Nevertheless, not all PTVs are pathogenic: fewer than 1,000 PTVs are classified as benign, while more than 10,000 remain annotated as VUS or as having conflicting interpretations.

Predictions of NMD are also imperfect. The first ∼150 nucleotides of the coding sequence, large exons (>400 nt), the last ∼55 nucleotides of the penultimate exon, and the final exon are predicted to be NMD-insensitive; however, approximately 30% of PTVs outside these regions still escape NMD.^[10–12]^. Establishing whether a PTV truly induces NMD can change the strength of ACMG/AMP PVS1 evidence^[13]^ and the variant classification. RNA-seq provides direct, empirical evidence by measuring transcriptional consequences and confirming or refuting NMD activity. In our cohort, 15 PTVs were analyzed: six showed no evidence of NMD, while nine clearly underwent decay. Notably, two PTVs located 70 and 100 nucleotides upstream of the final exon–exon junction, and one in the WARS2 pseudoexon, escaped NMD contrary to rule-based prediction. These findings indicate that a subset of PTVs in ClinVar may be misclassified due to erroneous NMD assumptions and should be re-evaluated using RNA-based evidence.

### Splicing disruption beyond canonical junctions

Classification of coding variants often relies on *in silico* predictions^[69]^. Although algorithms have improved and more than 40,000 missense variants are now annotated as (likely) pathogenic, many variants continue to be reclassified as new data emerge^[67, 70]^. Most coding variant predictors focus on protein structure or conservation but perform poorly for splicing effects, with average precision of only 1–2%^[71]^. Splice prediction algorithms such as SpliceAI, Pangolin, and MMSplice perform well for canonical junctions (accuracy 75–95%) but remain unreliable for near-splice, coding, or intronic regions^[71–73]^. Our analysis and previously published cohorts indicate that approximately 80% of pathogenic splice-altering variants are misclassified by such tools (Fig. 2C). Thus, both coding and non-coding variants can perturb RNA processing, and transcriptome profiling remains indispensable for mechanistic validation.

### A founder synonymous *ECHS1* variant in East Asian populations

We identified a recurrent synonymous *ECHS1* variant (c.489G>A, p.Pro163=) that induces partial exon 4 skipping, leading to decreased transcript levels and functional haploinsufficiency. This allele, enriched in East Asian populations (frequency 1.05%), represents a regional founder mutation. Its hypomorphic nature, still detected by RNAseq, explains the mild or late-onset presentations in compound heterozygotes and the apparently benign homozygous state observed in reference databases^[74]^. Parallel studies have reported this variant in 17 unrelated families from Japan, Samoa, and other Pacific populations, supporting a local founder effect^[74–76]^. Screening of our in-house Leigh syndrome cohort identified 13 additional cases, accounting for 42% of *ECHS1*-related diagnoses in Beijing. This finding reinforces the pathogenicity of the variant in trans with a known disease allele and highlights the importance of population-specific reference data in variant interpretation. Homozygous carriers of the *ECHS1* synonymous variant are functionally similar to individuals heterozygous for loss-of-function variants (LOF). Low constraint of *ECHS1* against LOF variants according to gnomAD explains the high frequency of homozygous carriers of this synonymous variant in East Asian populations. Importantly, *ECHS1* deficiency is a potentially treatable cause of Leigh syndrome, responsive to valine restriction and acetylcysteine supplementation^[47, 48]^.

### Deep intronic and structural variants revealed by RNA-seq

RNA-seq proved particularly effective in detecting pathogenic deep intronic variants invisible to WES and difficult to interpret from WGS. In two cases, pseudoexon creation in *WARS2* and *SUCLG1* produced premature stop codons and loss of function, illustrating how cryptic splice activation contributes to disease. RNA-seq–guided WGS also uncovered intragenic deletions in *PANK2* and *SERAC1* missed by WES, confirming that aberrant expression outliers can pinpoint underlying structural variants. Together, these examples highlight RNA-seq not only as a confirmatory tool but also as a discovery instrument that directs targeted reanalysis of genome data.

When splice defects lead to transcript degradation via NMD, they may manifest as expression outliers. Treatment of cultured cells with NMD inhibitors, such as cycloheximide or puromycin, can stabilize aberrant transcripts and facilitate their detection. However, a recent study by De Cock et al. (2025)^[77]^ demonstrated that while such treatment increases transcript coverage and refines splice characterization, it does not necessarily increase diagnostic yield.

### Broader implications and limitations

Integrating RNA-seq into clinical diagnostics shifts the focus from variant detection to variant interpretation. In our fibroblast dataset, 15,536 genes were expressed, representing two-thirds of all Mendelian disease genes, 90% of mitochondrial genes, and 92% of Leigh syndrome-associated genes, demonstrating that fibroblasts are a robust model for functional testing. However, gene expression is tissue-specific; genes restricted to the brain or muscle remain inaccessible in fibroblasts. Combining fibroblast transcriptomes with induced pluripotent stem cell–derived or neuron-specific RNA-seq may help overcome this limitation. To address tissue-specific expression, Hölzlwimmer et al. (2025) analyzed the GTEx dataset, which comprises transcriptomic and genomic data from 633 individuals across 49 tissues. They developed AbExp, a machine-learning-based approach that predicts tissue-specific gene expression by integrating genomic data with transcriptomes from clinically accessible tissues. In a dystonia cohort, AbExp-based prediction of brain gene expression using genomic data and fibroblast RNA-seq confirmed downregulation in the brain for genes already identified as expression outliers in fibroblasts. However, this approach did not reveal additional aberrant RNA events beyond those detectable in fibroblast cell lines, supporting the utility of fibroblast cell lines for functional studies in neurological disorders.

Despite the diagnostic efficacy of RNA-seq, many patients remained genetically unsolved, suggesting additional pathogenic mechanisms. In some mitochondrial disorders, *de novo* missense variants can be causative, as recently shown for a number of ATP synthase subunits^[78, 79]^; they can be inherited dominantly and with reduced penetrance or cause Leigh syndrome through digenic interactions between mtDNA and nuclear genes^[80]^. Rare combinations, such as biallelic *DNAJC30* variants plus additional pathogenic complex I variants, illustrate the diversity of inheritance models ^[81, 82]^. However, such events are extremely rare, and their identification requires statistical analysis of a large cohort of patients.

Future integration of long-read sequencing and complementary omics, including proteomics and methylomics, promises to further enhance diagnostic power. Proteomic studies have already demonstrated the ability to capture protein-destabilizing missense effects and reveal pathogenic variants not prioritized genomically^[83, 84]^. Understanding underexpression outliers can be challenging. Ozaki et al. (2024) required long-read WGS to identify a repeat expansion in the *NAXE* promoter as the cause of reduced expression in a patient with mitochondrial encephalopathy^[85]^. Moreover, epigenetic mechanisms can contribute to gene expression outliers, as exemplified by the discovery of promoter hypermethylation of *NDUFAF2* and its epigenetic silencing due to *MORC2* mutations in *trans,* associated with Leigh syndrome ^[86]^. Indeed, aberrant expression of *NDUFAF2* in two patients with Leigh syndrome within our cohort has been identified, which was caused by *MORC2* de novo mutations. A more non-specific *trans* effect on gene expression was recently reported in a patient with mitochondrial disease and pathogenic variants in *RNU4-2*. *RNU4-2* encodes a small nuclear RNA that plays a crucial role in the major spliceosome, which is the cellular machinery responsible for removing introns from pre-mRNA^[87]^. RNA-seq can also reveal systematic *trans*-splicing defects. By analysing excess intron retention outliers, Arriaga et al., (2025)^[88]^ identified four patients with biallelic variants in another RNA gene RNU4ATAC. Such examples highlight how RNA-seq captures both cis- and trans-acting transcriptomic perturbations and variant-to-function interpretation, broadening its diagnostic reach.

## Conclusion

Our results establish RNA-seq as a decisive functional complement to exome and genome sequencing in mitochondrial disease diagnostics. By revealing cryptic splicing defects, NMD escape, and non-coding variant effects, transcriptome profiling bridges the gap between genotype and phenotype, providing direct functional validation within a clinically relevant timeframe. Beyond its immediate diagnostic impact, the transcriptome analysis deepens mechanistic understanding of disease pathology and paves the way toward comprehensive, multi-omic precision medicine in Mendelian disorders.

## Data Availability

All data produced in the present study are available upon reasonable request to the authors

## Acknowledgements

We thank patients and their families for participation in this study. We would like to thank Editage (www.editage.cn) for English language editing. We thank René Santer for the critical reading of the manuscript.

